# Causal Effects of Corneal Traits on the risk of Keratoconus Using Genetic Instruments

**DOI:** 10.1101/2025.07.16.25331641

**Authors:** Bing Zhang

## Abstract

**Purpose:** To determine the causal effects of corneal traits on keratoconus (KC) using Mendelian randomization (MR) analysis.

**Design:** This is a two-sample Mendelian randomization (MR) study.

**Methods:** We performed two-sample MR analyses using European-ancestry genetic data. Genetic instruments for central corneal thickness (CCT), corneal hysteresis (CH), and corneal resistance factor (CRF) were analyzed against KC risk. Inverse variance weighted (IVW) regression served as the primary method to estimate causal effects.

**Results:** IVW analysis demonstrated that thicker CCT (per 1 mm) was causally associated with reduced KC risk (OR: 0.978, 95% CI: 0.961 to 0.995, P=0.01). Genetically predicted increases in right/left CH and right/left CRF (per 1 mmHg) were strongly inversely associated with KC (OR range: 0.165 to 0.198; all P<1.9×10⁻¹⁵). Sensitivity analyses confirmed robustness and directionality of these effects.

**Conclusion:** A causal effect between corneal traits and KC were indicated. Corneal biomechanical integrity, reflected by increased central corneal thickness, hysteresis, and resistance factor, causally protects against KC while established disease accelerates corneal degradation.

## INTRODUCTION

Keratoconus (KC) is a progressive corneal disorder characterized by thinning and conical protrusion of the cornea, leading to irregular astigmatism, visual impairment, and, in severe cases, the need for corneal transplantation.^1, 2^ The disease typically manifests during adolescence and young adulthood, significantly impacting patients’ quality of life and imposing a substantial economic burden on healthcare systems.^2–5^ Global prevalence estimates vary, but KC is recognized as a leading cause of corneal ectasia, with higher rates observed in certain ethnic populations.^6^ Despite advancements in diagnostic and therapeutic approaches, the deficiencies in screening methods remain prominent, underscoring the need for further research.

Identifying screening factors for KC is critical for early and targeted prevention. Corneal traits, including thickness and biomechanical parameters, can be measured non-invasively, holding potential as a population-level surveillance method. Although significant alterations in corneal traits (e.g., corneal thinning and reduced biomechanical integrity) are consistently observed in KC patients, these findings are predominantly interpreted as pathological consequences of the disease.^7, 8^ Crucially, there remains limited evidence on whether these traits have inherent value for screening at-risk individuals before clinical onset of disease. Generating such high-quality evidence traditionally requires large-scale, prospective cohort studies, which demand extended follow-up.

Mendelian randomization (MR), a genetic instrumental variable method, offers a powerful alternative by leveraging genetic variants associated with modifiable exposures (e.g., corneal traits) to infer causal relationships with outcomes (e.g., KC).^9^ Since genetic alleles are randomly assigned at conception, MR minimizes confounding and reverse causation biases inherent in observational studies. In this study, we apply a two-sample MR to elucidate the causal relationships between genetically predicted corneal traits and KC risk. This approach provides a novel pathway to investigate whether these traits play an etiological role in KC susceptibility, offering insights essential for refining risk stratification.

## METHODS

We retrieved the data of corneal traits from the MRC-IEU genome-wide association studies (GWAS) database (via https://gwas.mrcieu.ac.uk/) ^10, 11^, including central corneal thickness (CCT, ebi-a-GCST006366), right/left corneal hysteresis (RCH/LCH, ukb-b-11480/ukb-b-11650), and right/left corneal resistance factor (RCR/LCR, ukb-b-15178/ukb-b-4717). Additionally, we obtained data on KC from Release 12 of the FinnGen Project^12^, alongside a GWAS study (PMID: 33649486) that was analyzed in the sensitivity analysis ^13^.

This research utilized de-identified summary statistics from pre-ethically approved studies; thus, no additional institutional review board approval was required.

### Measurement of corneal traits

The central corneal thickness data utilized in this Mendelian randomization study were derived from a GWAS study of CCT cohorts from the International Glaucoma Genetics consortium using imputation based on the 1000 Genomes Project (PMID: 29760442) ^14^. The 1000 Genomes Project, initiated in 2008, aims to generate a comprehensive catalogue of human genetic variation through international collaboration.^15^ This GWAS study comprehensively analyzed CCT measurements from 17,803 individuals of European descent across 14 independent cohorts. The European ancestry-specific meta-analysis within this GWAS study served as the foundation for our selection of genome-wide significant CCT loci.

Corneal hysteresis and resistance factor were using ocular response analyzer (ORA, Reichert, Inc., Depew, NY) in UK Biobank. The UK Biobank is a large-scale biomedical database with de-identified genetic, lifestyle, and health data from about 500,000 UK participants, which was approved by the North West Multi-centre Research Ethics Committee (Ref No. 06/MRE08/65). In the eye subcohort, CH and CR was measured following a predetermined protocol, excluding participants who had undergone eye surgery within the last 4 weeks; measurements were initially taken in the right eye and performed only once per eye, with a second attempt if the participant blinked during the procedure.^16^ Of the participants with corneal biomechanical data, European descents with GWAS data were analyzed and the detailed information on the GWAS analyses is available in the published literature.^17^ <u>Measurement of KC</u>

KC data were obtained from Release 12 of the FinnGen Project. The FinnGen Project is a large-scale research initiative that combines genotype data from Finnish biobanks with digital health records from Finnish health registries to understand the genetic basis of diseases, with its study protocol approved by the Coordinating Ethics Committee of the Hospital District of Helsinki and Uusimaa (Ref. No. HUS/990/2017) ^12^. In the FinnGen project, KC is defined based on diagnostic codes from hospital discharge records and cause of death registries. For the R12 GWAS, there are 878 cases and 473,095 controls enrolled.

KC data from a GWAS meta-analysis (PMID: 33649486), comprising 4,669 cases and 116,547 controls were applied in the sensitivity analysis^13^. The cases were from the 1000 Genomes Project and rigorously diagnosed based on characteristic signs, and confirmed with Orbscan/Pentacam, while the controls were from UK Biobank without any ICD9 or ICD10 code for any corneal disease. Individuals with prior bilateral keratoplasty for KC were included while patients with syndromic associations (e.g., Down syndrome) or other corneal dystrophies/ectasias were excluded. However, due to partial sample overlap (cases with CCT data of 1000 Genomes, controls with corneal biomechanics data of UK Biobank) and non-exclusive European ancestry (89%), this KC dataset violates key two-sample MR assumptions (sample independence and population homogeneity). Thus, it was used solely for sensitivity analysis to verify the robustness of the FinnGen-based findings.

### Mendelian Randomization

We conducted two-sample MR analyses using the TwoSampleMR package in R.^18^ We selected single nucleotide polymorphisms (SNPs) that reached genome-wide significance (p < 5×10⁻⁸) for each corneal trait. SNPs were pruned for linkage disequilibrium at R² < 0.001 over a 10000 KB window, selecting the SNP with the lowest p-value within each locus.

Primary causal estimates were derived from inverse-variance weighted (IVW) regression with multiplicative random effects, which served as the principal analytical method due to its recognized effectiveness.^19^ Supplementary analyses employed weighted median estimation, MR-Egger regression, and simple and weighted mode estimation. Statistical significance was primarily assessed using inverse-variance weighted (IVW) analysis, with a Bonferroni-corrected significance threshold of P < 0.01 applied for multiple comparisons (n corneal traits=5).

### Sensitivity Analysis

Sensitivity analyses were conducted to validate result robustness^20^: instrument strength was evaluated via F-statistic to assess variant relevance as IVs, variant heterogeneity was tested using Cochran’s Q test, potential bias from horizontal pleiotropy was examined by MR-Egger regression, and outlier influence was evaluated through leave-one-out sensitivity analysis—only the cross-validation result for the SNP exerting the most substantial impact on the outcomes (yielding the highest P-value upon removal) were reported. The Steiger directionality test was performed to verify the correct causal direction between exposure and outcome.^18^ Despite its limitations, we replicated the MR analysis using the KC dataset (PMID: 33649486) to find consistent results with the FinnGen cohort.

## RESULTS

### Genetic Data Resources and Instrument Selection

This Mendelian randomization study utilized summary-level genome-wide association statistics from European-ancestry cohorts to evaluate the causal influence of corneal traits on keratoconus (KC). As detailed in Table 1, CCT data originated from a meta-analysis of 17,803 individuals (GWAS ID: ebi-a-GCST006366), while corneal hysteresis (right: RCH; left: LCH) and resistance factors (right: RCR; left: LCR) were derived from UK Biobank (97,653-97,465 participants). Primary KC outcome data were obtained from FinnGen R12 (878 cases and 473,095 controls). For sensitivity verification, an independent KC dataset (PMID: 33649486 with 4,669 cases and 116,547 controls) was incorporated despite known limitations of partial sample overlap.

**Table 1.** Summary Information on the Genome-Wide Association Study (GWAS) Database in This Study.

| **Trait** | **Trait Abbr.** | **GWAS ID** | **Data Source** | **Sample** | **Sample Size** | **Population** |
| --- | --- | --- | --- | --- | --- | --- |
| Central Corneal thickness | CCT | ebi-a-GCST006366 | MRC-IEU | Meta-analysis | 17,803 | European |
| Corneal hysteresis (right) | RCH | ukb-b-11480 | MRC-IEU | UK Biobank | 97,653 | European |
| Corneal hysteresis (left) | LCH | ukb-b-11650 | MRC-IEU | UK Biobank | 97,465 | European |
| Corneal resistance factor (right) | RCR | ukb-b-15178 | MRC-IEU | UK Biobank | 97,653 | European |
| Corneal resistance factor (left) | LCR | ukb-b-4717 | MRC-IEU | UK Biobank | 97,465 | European |
| Keratoconus | KC | - | FinnGen | FinnGen R12 | 473,973<br>(878 cases) | European |
| Keratoconus<br>(for Sensitivity Analysis) | KC | - | PubMed<br>(33649486) | Cases: 1000 Genomes<br>Controls: UK Biobank | 121,216<br>(4,669 cases) | European (89%) |

### Causal Effects of Corneal Traits on Keratoconus

IVW regression analyses demonstrated statistically significant protective effects for all corneal parameters against KC development (Table 2). Each 1-mm increase in CCT reduced KC risk by 2.2% (OR 0.978, 95% CI: 0.961-0.995; P = 0.01). Corneal biomechanical traits exhibited significant effects: per 1-mmHg increment, RCH decreased KC risk by 83.5% (OR 0.165, 95% CI: 0.112-0.241; P = 1.9×10⁻²⁰), LCH by 83.9% (OR 0.161, 95% CI: 0.103-0.253; P = 1.9×10⁻¹⁵), RCR by 81.7% (OR 0.183, 95% CI: 0.135-0.247; P = 3.3×10⁻²⁸), and LCR by 80.2% (OR 0.198, 95% CI: 0.144-0.274; P = 1.2×10⁻²²). Consistency across MR methods was visually confirmed in Figure 1, where scatterplots of genetic associations show regression lines from five MR methods converging toward protective effects for all corneal parameters.

**Figure 1:**
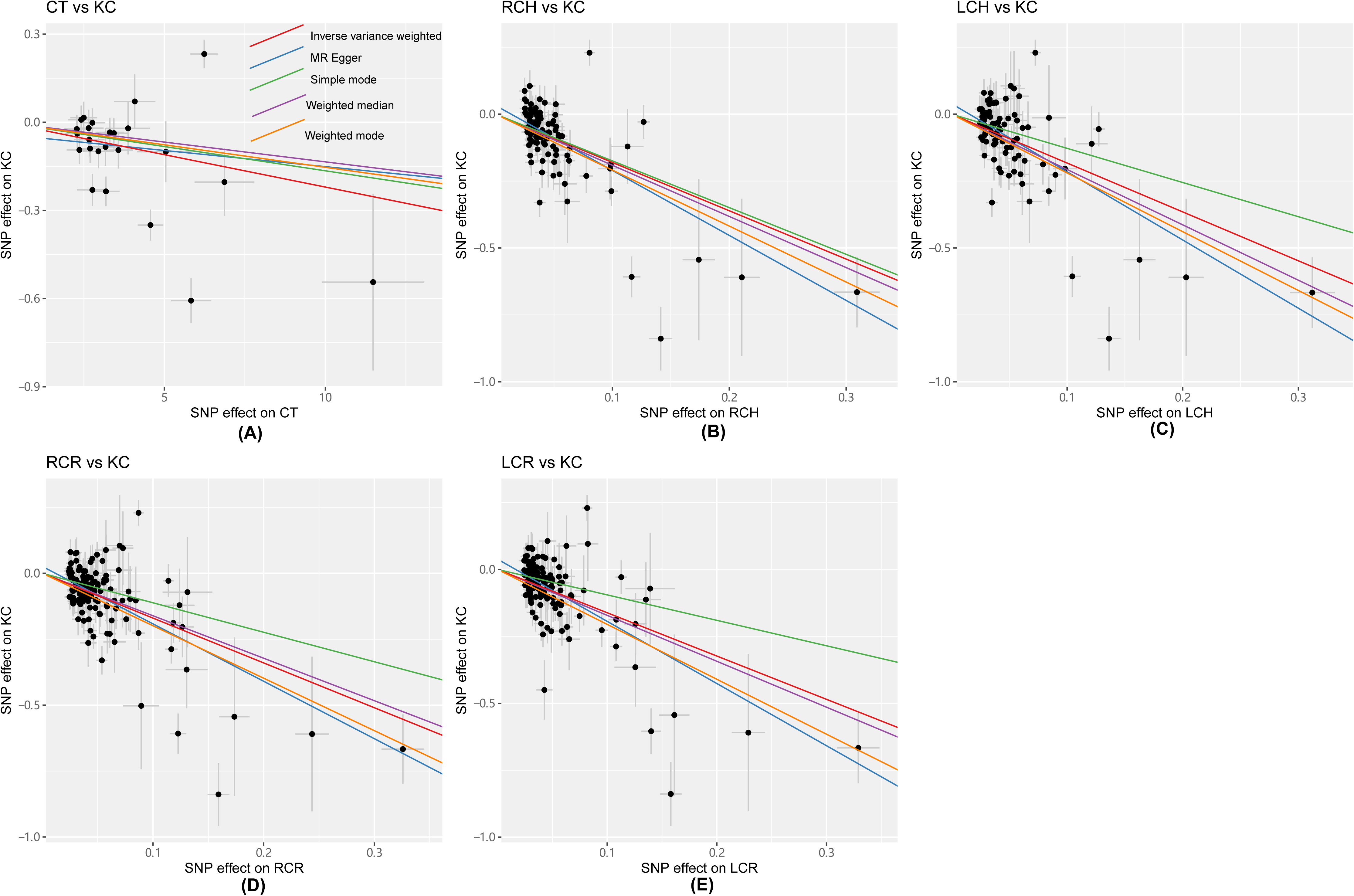
Scatterplots depicting genetic associations between corneal parameters and KC risk. Regression lines from five MR methods illustrate consistent causal directions. CCT = central corneal thickness; RCH = corneal hysteresis (right); LCH = corneal hysteresis (left); RCR = corneal resistance factor (right); LCR = corneal resistance factor (left).

**Table 2.** Mendelian Randomization Analysis of Causal Effects of Corneal Parameters on Keratoconus.^†^.

| **Exposure (Abbr.)** | **No SNPs** | **Method** | **OR (95% CI)** | **P value** |
| --- | --- | --- | --- | --- |
| CCT, per mm | 24 | MR Egger | 0.989(0.939, 1.042) | 0.68 |
|  |  | Weighted median | 0.987(0.975, 0.999) | 0.03 |
|  |  | **<u>IVW</u>** | **<u>0.978(0.961, 0.995)</u>** | **<u>0.01</u>** |
|  |  | Simple mode | 0.984(0.966, 1.002) | 0.09 |
|  |  | Weighted mode | 0.985(0.961, 1.009) | 0.22 |
| RCH, per mmHg | 105 | MR Egger | 0.088(0.039, 0.2) | 6.6E-08 |
|  |  | Weighted median | 0.148(0.097, 0.226) | 1.1E-18 |
|  |  | **<u>IVW</u>** | **<u>0.165(0.112, 0.241)</u>** | **<u>1.9E-20</u>** |
|  |  | Simple mode | 0.175(0.065, 0.47) | 8E-4 |
|  |  | Weighted mode | 0.124(0.06, 0.256) | 1.6E-07 |
| LCH, per mmHg | 88 | MR Egger | 0.078(0.031, 0.199) | 7.3E-07 |
|  |  | Weighted median | 0.126(0.077, 0.207) | 2E-16 |
|  |  | **<u>IVW</u>** | **<u>0.161(0.103, 0.253)</u>** | **<u>1.9E-15</u>** |
|  |  | Simple mode | 0.279(0.084, 0.923) | 0.04 |
|  |  | Weighted mode | 0.111(0.051, 0.242) | 3.3E-07 |
| RCR, per mmHg | 140 | MR Egger | 0.113(0.059, 0.218) | 1.1E-09 |
|  |  | Weighted median | 0.2(0.138, 0.29) | 2.2E-17 |
|  |  | **<u>IVW</u>** | **<u>0.183(0.135, 0.247)</u>** | **<u>3.3E-28</u>** |
|  |  | Simple mode | 0.327(0.126, 0.848) | 0.02 |
|  |  | Weighted mode | 0.137(0.078, 0.241) | 1.9E-10 |
| LCR, per mmHg | 124 | MR Egger | 0.098(0.05, 0.192) | 4.5E-10 |
|  |  | Weighted median | 0.18(0.122, 0.265) | 5.2E-18 |
|  |  | **<u>IVW</u>** | **<u>0.198(0.144, 0.274)</u>** | **<u>1.2E-22</u>** |
|  |  | Simple mode | 0.387(0.134, 1.112) | 0.08 |
|  |  | Weighted mode | 0.129(0.069, 0.24) | 2.5E-09 |

<sup>†</sup> Abbreviations: OR = odds ratio; CI = confidence interval; IVW= Inverse variance weighted; CCT = central corneal thickness; RCH = corneal hysteresis (right); LCH = corneal hysteresis (left); RCR = corneal resistance factor (right); LCR = corneal resistance factor (left).

### Sensitivity Analysis: Methodological Validation

Comprehensive sensitivity analyses confirmed instrument validity and result robustness (Table 3). Instrument strength was supported by mean F-statistics >53.3 (range: 53.3 for CCT to 77.6 for RCR), exceeding the critical threshold of 10. Significant heterogeneity was detected (Cochran’s Q >153, P <3.4×10⁻²¹), necessitating random-effects IVW models. Horizontal pleiotropy was negligible for most traits (MR-Egger intercept P >0.089), though LCR showed marginal evidence (P=0.021, which is negligible at Bonferroni-corrected threshold of 0.01). Leave-one-out sensitivity analysis confirmed robustness for corneal biomechanical traits: removal of the most influential SNP preserved strong statistical significance (P₁ = 3.3×10⁻¹⁸ to 1.6×10⁻¹³) with minimal effect estimate changes (|Δβ/β₀| ≤ 2.0%). For central corneal thickness, however, SNP exclusion reduced significance from P₀ = 0.01 to P₁ = 0.05 and altered the IVW estimate by 23.6%, indicating relative instability. Steiger directionality tests confirmed correct causal ordering (P <4×10⁻²⁵⁴).

**Table 3.** Methodological assessment of the Mendelian randomization analyses: Instrument strength, heterogeneity, horizontal pleiotropy, leave-one-out and directionality analysis.^†^.

| Analysis | Instrument Strength | Heterogeneity |  | Horizontal Pleiotropy |  | Leave-One-Out Analysis* |  |  |  |  |  | Directionality |
| --- | --- | --- | --- | --- | --- | --- | --- | --- | --- | --- | --- | --- |
| | (mean F value) | (Q value) | (P value) | (Intercept) | (P value) | $\beta_0$ | $P_0$ | $\beta_1$ | $P_1$ | $\Delta\beta$ | $\vert \Delta\beta/\beta_0\vert $ % | Steiger P value |
| CCT -> KC | 53.3 | 153.0 | 3.4E-21 | -0.0409 | 0.661 | -0.022 | 0.01 | -0.017 | 0.05 | 0.005 | 23.6 | 4E-254 |
| RCH -> KC | 72.0 | 266.1 | 3.6E-16 | 0.032459 | 0.095 | -1.805 | 1.9E-20 | -1.774 | 3.3E-18 | 0.031 | 1.7 | <1E-300 |
| LCH -> KC | 74.0 | 262.9 | 1.4E-19 | 0.039401 | 0.089 | -1.826 | 1.8E-15 | -1.792 | 1.6E-13 | 0.034 | 1.8 | <1E-300 |
| RCR -> KC | 77.6 | 308.3 | 7.3E-15 | 0.025823 | 0.108 | -1.700 | 3.3E-28 | -1.675 | 1.2E-25 | 0.024 | 1.4 | <1E-300 |
| LCR -> KC | 76.2 | 278.9 | 4.0E-14 | 0.038263 | 0.021 | -1.617 | 1.2E-22 | -1.584 | 3.2E-20 | 0.033 | 2.0 | <1E-300 |

<sup>†</sup> Abbreviations: CCT = central corneal thickness; RCH = corneal hysteresis (right); LCH = corneal hysteresis (left); RCR = corneal resistance factor (right); LCR = corneal resistance factor (left).
\* Leave-one-out cross-validation was performed on the single instrumental variable exerting the most substantial impact on the results, the one generating the highest P value upon removal. $\beta_0/P_0$ : Inverse-variance weighted (IVW) estimate and its statistical significance using all SNPs. $\beta_1/P_1$ : IVW estimate and its statistical significance after removing the key SNP. $\Delta\beta$ : Absolute change in effect estimate caused by SNP removal. $(\Delta\beta/\beta_0)\%$ : percentage change of $\Delta\beta/\beta_0$ .

### Sensitivity Analysis: with Alternative KC Dataset

MR analyses replicated in the independent KC cohort yielded highly concordant results (Table 4). IVW estimates confirmed protective effects: CCT (OR 0.978, 95% CI: 0.963-0.994; P=0.006), RCH (OR 0.190, 95% CI: 0.130-0.277; P=7.18×10⁻¹⁸), LCH (OR 0.183, 95% CI: 0.120-0.280; P=4.40×10⁻¹⁵), RCR (OR 0.224, 95% CI: 0.166-0.302; P=6.48×10⁻²³), and LCR (OR 0.201, 95% CI: 0.142-0.285; P=1.88×10⁻¹⁹). Weighted median estimates provided additional confirmation (all P<9.56×10⁻¹⁵), establishing result stability despite methodological constraints in this dataset.

**Table 4.** Sensitivity Analysis of the Mendelian Randomization of Corneal Parameters on Keratoconus.^†^.

| Exposure (Abbr.) | No SNPs | Method | OR (95% CI) | P value |
| --- | --- | --- | --- | --- |
| CCT, per mm | 22 | MR Egger | 0.999(0.951, 1.05) | 0.964 |
|  |  | Weighted median | 0.986(0.979, 0.992) | 3.46E-05 |
|  |  | **IVW** | **0.978(0.963, 0.994)** | **0.006** |
|  |  | Simple mode | 0.986(0.977, 0.994) | 0.004 |
|  |  | Weighted mode | 0.986(0.978, 0.994) | 0.003 |
| RCH, per mmHg | 86 | MR Egger | 0.192(0.073, 0.504) | 0.001 |
|  |  | Weighted median | 0.272(0.202, 0.368) | 1.80E-17 |
|  |  | **IVW** | **0.19(0.13, 0.277)** | **7.18E-18** |
|  |  | Simple mode | 0.306(0.127, 0.739) | 0.01 |
|  |  | Weighted mode | 0.385(0.23, 0.645) | 0.0005 |
| LCH, per mmHg | 76 | MR Egger | 0.151(0.053, 0.432) | 0.0007 |
|  |  | Weighted median | 0.268(0.192, 0.374) | 9.56E-15 |
|  |  | **IVW** | **0.183(0.12, 0.28)** | **4.40E-15** |
|  |  | Simple mode | 0.436(0.132, 1.437) | 0.177 |
|  |  | Weighted mode | 0.412(0.193, 0.883) | 0.026 |
| RCR, per mmHg | 123 | MR Egger | 0.203(0.095, 0.434) | 7.32E-05 |
|  |  | Weighted median | 0.348(0.272, 0.445) | 5.17E-17 |
|  |  | **IVW** | **0.224(0.166, 0.302)** | **6.48E-23** |
|  |  | Simple mode | 0.502(0.23, 1.097) | 0.086 |
|  |  | Weighted mode | 0.430(0.262, 0.705) | 0.001 |
| LCR, per mmHg | 99 | MR Egger | 0.128(0.054, 0.303) | 9.42E-06 |
|  |  | Weighted median | 0.316(0.238, 0.42) | 1.72E-15 |
|  |  | **IVW** | **0.201(0.142, 0.285)** | **1.88E-19** |
|  |  | Simple mode | 0.410(0.197, 0.853) | 0.019 |
|  |  | Weighted mode | 0.401(0.211, 0.760) | 0.006 |

Abbreviations: OR = odds ratio; CI = confidence interval; IVW= Inverse variance weighted; CCT = central corneal thickness; RCH = corneal hysteresis (right); LCH = corneal hysteresis (left); RCR = corneal resistance factor (right); LCR = corneal resistance factor (left).

## DISCUSSION

This Mendelian randomization study establishes robust genetic evidence that corneal structural and biomechanical traits, specifically central corneal thickness, hysteresis, and resistance factor, exert causal effects on keratoconus. Unlike observational studies constrained by confounding and reverse causation, the genetic approach demonstrates that intrinsically thicker corneas and enhanced biomechanical integrity directly reduce KC susceptibility.

### Corneal Traits on Keratoconus

The primary IVW analyses provide compelling genetic evidence that corneal traits directly influence KC susceptibility. Specifically, each 1-mm increase in central corneal thickness (CCT) was associated with a 2.2% reduction in KC risk, whereas corneal hysteresis and resistance factors exhibited pronounced protective effects with each 1-mmHg increment conferring over an 80% risk reduction. While MR estimates may be susceptible to inflated effect sizes, the exceptionally strong and significant ORs observed in this study suggest substantial clinical applicability.^21^ These findings, consistent across multiple MR methods, reclassify corneal traits as risk determinants rather than passive disease markers. Unlike observational studies, which have often reported observational associations between altered corneal traits and keratoconus,^22, 23^ this study demonstrates that genetically determined corneal thickness and biomechanical properties act as risk factors against KC pathogenesis by leveraging genetic instruments. Notably, thicker corneas and enhanced biomechanical integrity directly reduce KC susceptibility. This profound protective effect aligns well with established collagen cross-linking theories^2^; biomechanically, the increased corneal biomechanical integrity can effectively prevent biomechanical failure and the subsequent ectatic progression characteristic of KC.

Comprehensive sensitivity analyses validated result robustness. Methodological validation (Table 3) confirmed instrument strength, revealed no significant heterogeneity or horizontal pleiotropy, and demonstrated consistent directionality across analyses. While leave-one-out sensitivity analysis indicated relative instability in CCT estimates (P=0.05), hysteresis and resistance factor results remained highly stable. Analysis in an alternative keratoconus dataset yielded consistent protective effects, with effect sizes (ORs) comparable to primary analyses with FinnGen R12 dataset. Crucially, this independent validation confirmed the stability of CCT’s protective association. These findings establish corneal biomechanical properties as fundamental determinants in keratoconus pathogenesis.

### Clinical and Research Implications

Our study provides genetic validation for integrating corneal traits into KC risk stratification. The findings indicate measuring CCT and corneal biomechanical traits, such as CH and CR, during routine ophthalmological exams or community-based screenings could identify high-risk individuals at subclinical stages, enabling timely monitoring or early intervention. Future research should prioritize developing standardized corneal trait-based screening protocols for KC, establishing longitudinal cohorts to validate the prognostic value of corneal traits for disease progression, and elucidating the mechanistic pathways by which corneal traits directly influence KC susceptibility. Additionally, validating these approaches in multi-ethnic populations and optimizing risk prediction models are essential to translate these research advances into effective clinical practice.

### Limitation

While leveraging large-scale genetic consortia and rigorous sensitivity analyses, several limitations merit acknowledgment. First, the exclusive European ancestry of GWAS data restricts generalizability; validation in diverse populations is essential. Second, although MR-Egger intercepts indicated minimal horizontal pleiotropy (Table 3), residual confounding cannot be fully excluded. Third, while establishing genetic causality, this study does not delineate specific biological mediators.

## Conclusion

In summary, this study provides robust evidence that genetically determined increases in central corneal thickness, hysteresis, and resistance factor confer significant protection against keratoconus. The consistency of these effects across methodological approaches and sensitivity analysis. These findings advocate for clinical adoption of corneal thickness and biomechanical phenotyping in screening protocols and underscore the promise of early interventions targeting keratoconus.

## Data Availability

All data produced in the present study are available upon reasonable request to the authors

## Abbreviations

CCT: Central Corneal Thickness
CH: Corneal Hysteresis
CI: Confidence Interval
CRF: Corneal Resistance Factor
GWAS: Genome-Wide Association Study
IVW: Inverse Variance Weighted KC, Keratoconus
RCR: Right Corneal Resistance Factor
SNP: Single Nucleotide Polymorphism
LCH: Left Corneal Hysteresis
LCR: Left Corneal Resistance Factor
MR: Mendelian Randomization
MRC-IEU: Medical Research Council Integrative Epidemiology Unit
ORA: Ocular Response Analyzer OR, Odds Ratio
RCH: Right Corneal Hysteresis

## Meeting Presentation

None.

## Data availability

All data used in this study were obtained from publicly available databases. Genomic and phenotypic data for corneal traits (corneal thickness, corneal hysteresis, and corneal resistance factor) were derived from the MRC-IEU OpenGWAS database (https://gwas.mrcieu.ac.uk/). The keratoconus dataset was sourced from FinnGen R12 (via https://storage.googleapis.com/finngen-public-data-r12/summary_stats/release/finngen_R12_H7_CORNEALDEFORM.gz) and a published meta-analysis (PubMed ID: 33649486, via https://pmc.ncbi.nlm.nih.gov/articles/instance/7921564/bin/42003_2021_1784_MOESM4_ESM.txt”). The analytical code used for Mendelian randomization and statistical analyses is available from the corresponding author upon reasonable request.

